# Phylogeography and reassortment patterns of human influenza A viruses in sub-Saharan Africa

**DOI:** 10.1101/2024.01.07.24300955

**Authors:** D. Collins Owuor, Zaydah R. de Laurent, John W. Oketch, Nickson Murunga, James R. Otieno, Sandra S. Chaves, D. James Nokes, Charles N. Agoti

## Abstract

**Background:** The role of sub-Saharan Africa in the global spread of influenza viruses remains unclear due to insufficient spatiotemporal sequence data.

**Methods:** Here, we analyzed 222 codon-complete sequences of influenza A viruses (IAVs) sampled between 2011 and 2013 from five countries across sub-Saharan Africa (Kenya, Zambia, Mali, Gambia, and South Africa); these genomes were compared with 1,209 contemporaneous global genomes using phylogeographical approaches.

**Results:** The spread of influenza in sub-Saharan Africa was characterized by (i) multiple introductions of IAVs into the region over consecutive influenza seasons, with viral importations originating from multiple global geographical regions, some of which persisted in circulation as intra-subtype reassortants for multiple seasons, (ii) virus transfer between sub-Saharan African countries, and (iii) virus export from sub-Saharan Africa to other geographical regions.

**Conclusion:** Despite sparse data from influenza surveillance in sub-Saharan Africa, our findings support the notion that influenza viruses persist as temporally structured migrating metapopulations in which new virus strains can emerge in any geographical region, including in sub-Saharan Africa; these lineages may have been capable of dissemination to other continents through a globally migrating virus population. Further knowledge of the viral lineages that circulate within understudied sub-Saharan Africa regions is required to inform vaccination strategies in those regions.

## Introduction

The rapid and widespread global circulation of influenza A(H1N1)pdm09 virus in 2009 [1–4] and severe acute respiratory syndrome coronavirus 2 (SARS-CoV-2) in 2020 [5–9] demonstrate how respiratory viruses can quickly spread globally following their emergence. Influenza A viruses (IAV) and novel coronaviruses remain a major public health threat with potential to cause pandemics resulting in economic losses, social lockdowns, and millions of deaths [10–13]. Therefore, global surveillance efforts to monitor these pathogens through various initiatives, for example, the Global Influenza Surveillance and Response System (GISRS) [14] remain justified to improve understanding of their global transmission networks and dynamics for appropriate interventions.

The global surveillance of influenza through GISRS has resulted in the generation of geographically and temporally extensive virus sequence data, which has provided a unique opportunity to investigate the global spread of influenza viruses [15–20]. Three key hypotheses have been previously proposed to explain how influenza viruses spread globally. First, the “source-sink” model, whereby East and Southeast (E-SE) Asia represent a global source population of novel influenza virus strains, while temperate regions represent ecological sinks [15, 16, 18]. Second, influenza viruses occur as temporally migrating metapopulations, whereby new virus strains can emerge in any geographical region, with the location of the source population changing from season-to-season [17]. Third, the global patterns of spread of seasonal influenza viruses have been shown to vary with the rate of genetic and antigenic evolution of different influenza virus types and subtypes [19]. However, despite documented high disease burden [21–23], the role of sub-Saharan African countries in the global spread of influenza viruses remains unclear due to insufficient surveillance and sequence data from these regions [22, 24].

To improve understanding of IAV dispersal pathways in sub-Saharan Africa and the context of locally observed virus diversity, we studied the spread of A(H1N1)pdm09 and A(H3N2) viruses in the region using phylogeographical methods based on codon-complete sequences from samples obtained from a multi-country surveillance involving five sub-Saharan African countries. We then compared these data with sequences that were available from other countries in sub-Saharan Africa and around the globe.

## Materials and Methods

### Sample sources and molecular screening

The samples analysed here were collected under three distinct studies: (a) the Pneumonia Etiology Research for Child Health (PERCH) study, which enrolled children aged between 1-59 months admitted to hospital with severe or very severe pneumonia in surveillance sites in Kenya, Zambia, Mali, Gambia, and South Africa [25–27] from August 2011 through January 2014; (b) a Kenya-wide surveillance of influenza viruses in Kenya among patients of any age hospitalized with severe acute respiratory illness (SARI) [28] from January 2011 through December 2013; and (c) inpatient paediatric (1-59 months) pneumonia surveillance of influenza viruses in Kilifi County and Referral Hospital (KCH) [29] from January 2011 through December 2013. Detailed descriptions of the study sites and population, sample collection, transportation, storage, and processing have been described elsewhere; PERCH study [25, 27, 30], the countrywide surveillance study [28], and the inpatient paediatric pneumonia surveillance study [29]. For the PERCH study, a total of 138 IAV positive specimens were analysed from five surveillance sites from five countries, **Figure S1A**: Kenya, 27; South Africa, 40; Zambia, 27; The Gambia, 22; and Mali, 22. A further 106 A(H1N1)pdm09 and 16 A(H3N2) virus sequences were available from the countrywide surveillance study and the paediatric KCH study, respectively.

### RNA extraction and multi-segment real-time PCR (M-RTPCR) for IAV

Viral RNA extraction and M-RTPCR was undertaken as previously described [29]. Briefly, we performed viral nucleic acid extraction from IAV positive samples (Cycle threshold (Ct) <35.0) using the QIAamp Viral RNA Mini Kit (Qiagen). We reverse transcribed ribonucleic acid (RNA) and amplified the codon-complete region of influenza in a single M-RTPCR using the Uni/Inf primer set [31] in 25 μL PCR reactions. We evaluated successful amplification by running the M-RTPCR amplicons on 2% agarose gel and visualized the gels looking for expected bands on a UV transilluminator after staining with RedSafe Nucleic Acid Staining solution (iNtRON Biotechnology Inc.,).

### IAV codon-complete sequencing and genome assembly

Following M-RTPCR, we purified, quantitated and normalized amplicons to 0.2 ng/μL as described previously [29]. Briefly, the amplicons were purified with 1X AMPure XP beads (Beckman Coulter Inc.,), quantified with Quant-iT dsDNA High Sensitivity Assay (Invitrogen), and normalized to 0.2 ng/μL. Indexed paired end libraries were generated from 2.5 μL of 0.2 ng/μL amplicon pool using Nextera XT Sample Preparation Kit (Illumina) following the manufacturer’s protocol. Amplified libraries were purified using 0.8X AMPure XP beads, quantitated using Quant-iT dsDNA High Sensitivity Assay (Invitrogen), and evaluated for fragment size in the Agilent 2100 BioAnalyzer System using the Agilent High Sensitivity DNA Kit (Agilent Technologies). Libraries were then diluted to 2nM in preparation for pooling and denaturation for running on the Illumina MiSeq (Illumina). Pooled libraries were NaOH denatured, diluted to 12.5 pM and sequenced on the Illumina MiSeq using 2 x 250 bp paired end reads with the MiSeq v2 500 cycle kit (Illumina). Five percent Phi-X (Illumina) spike-in was added to the libraries to increase library diversity by creating a more diverse set of library clusters. We carried out contiguous nucleotide sequence assembly from the sequence data using the FLU module of the Iterative Refinement Meta-Assembler (IRMA) [32] using IRMA default settings. All generated sequence data were deposited in the Global Initiative on Sharing All Influenza Data (GISAID) EpiFlu^TM^ database (https://platform.gisaid.org/epi3/cfrontend) under the accession numbers EPI_ISL_509524-EPI_ISL_509526, EPI_ISL_509564-EPI_ISL_509566, EPI_ISL_509655-EPI_ISL_509669, EPI_ISL_509687, EPI_ISL_510040-EPI_ISL_510043, EPI_ISL_510078-EPI_ISL_510080, EPI_ISL_510102, EPI_ISL_510152-EPI_ISL_510159, EPI_ISL_509025-EPI_ISL_509058, EPI_ISL_509396-EPI_ISL_509411, and EPI_ISL_511774-EPI_ISL_511804.

### Collation of contemporaneous global sequence dataset

Global comparison datasets for influenza A(H1N1)pdm09 and A(H3N2) viruses were retrieved from the GISAID EpiFlu^TM^ database (https://platform.gisaid.org/epi3/cfrontend; accessed 20 March 2022). The datasets were prepared to determine the relatedness of the viruses in this report to those circulating around the world thus understand their global context. Only codon-complete full-length genome sequences sampled between January 2010 and December 2013 were included to improve the phylogenetic resolution of the analyses and investigate reassortment patterns. For A(H1N1)pdm09 virus, the final dataset of 460 global sequences sampled from January 2010 to December 2013 was available (numbers in parenthesis indicate number of sequences): Africa (13); Asia (148); Europe (82); North America (170); South America (2); Oceania (39). For A(H3N2) virus, the final dataset of 749 global sequences sampled from January 2010 to December 2013 was available (numbers in parenthesis indicate number of sequences): Africa (7); Asia (196); Europe (38); North America (178); South America (199); Oceania (138). The accession numbers for the global genome sequences are available in the study’s GitHub repository, (https://github.com/DCollinsOwuor/Phylogeography-and-reassortment-patterns-of-human-influenza-A-viruses-in-sub-Saharan-Africa).

### Phylogenetic analysis

We aligned and translated consensus nucleotide sequences in AliView v1.26 [33] and concatenated individual gene segments into codon-complete genomes using SequenceMatrix v16.0.1 [34]. We constructed codon-complete gene segment and concatenated genome phylogenetic trees of A(H1N1)pdm09 and A(H3N2) viruses with maximum-likelihood (ML) and bootstrap analysis of 1,000 replicates. We inferred the best-fit nucleotide substitution models using IQ-TREE v1.6.11 [35, 36] and implemented those chosen by the Bayesian Information Criterion for each concatenated virus genome. We visualized and annotated the phylogenetic trees using ggtree R package v2.3.5 [37] and used the full-length HA sequences of all virus sequences generated from the PERCH, countrywide surveillance, and inpatient paediatric pneumonia surveillance study to characterize A(H1N1)pdm09 and A(H3N2) viruses into clades using PhyCLIP v2.0 [38] and IAV clade representative strains.

### Reassortment analysis

We used tanglegrams of the time-resolved trees from TreeTime v0.8.5 [39] to visualize the phylogenetic relationships between individual codon-complete gene segments of A(H1N1)pdm09 and A(H3N2) viruses using dendextend R package v1.15.2 [40] and explored the pairwise phylogenetic congruence between IAV gene segments to identify intra-subtype reassortment. Tanglegrams are sets of two trees representing different segments of IAVs, with tips from matching viruses connected by lines. We compared topologies of tanglegram trees that matched the phylogeny of HA segment with that of other segments, coloring tanglegram twines based on observed reassortment patterns.

We then estimated the reassortment rates (the number of reassortment events per lineage per year (events/lineage/year)) among the PERCH study and sub-Saharan African IAVs using a coalescent reassortant constant population model (CoalRe) [41] in BEAST2 v.2.6.6 (https://www.beast2.org/) with parameters: GTR + I + G4 substitution model, strict clock, prior for reassortment rate = exponential with mean 0.25, chain length of 500 million, and 10 per cent burn-in. We then used the Graph-incompatibility-based Reassortment Finder (GiRaF) tool [42] to computationally assess reassortment among IAVs. Our GiRaF runs, that computationally assessed reassortments, converged with an average standard deviation of split frequency < 0.01, tree length estimated sample size (ESS) value > 100, tree length potential scale reduction factors (PSRF) of 1.0, and a maximum split frequency PSRF of 1.00–1.002 (**Tables 1** and **2**). We manually discarded the first 25 percent of trees from each of the two runs (burn-in), then combined and processed the remaining trees. Only reassortment events with a high confidence value (≥0.70) and reassortant sets predicted in at least three of twenty-eight gene pairwise comparisons, as recommended by GiRaF developers were reported. We ran two simultaneous runs of MrBayes v3.2.7 [43] on the datasets (A(H1N1)pdm09 viruses, 200 million generations; A(H3N2) viruses, 100 million generations) under a GTR+I+Γ substitution model and sampled trees at every 200,000th generation.

**Table 1.**
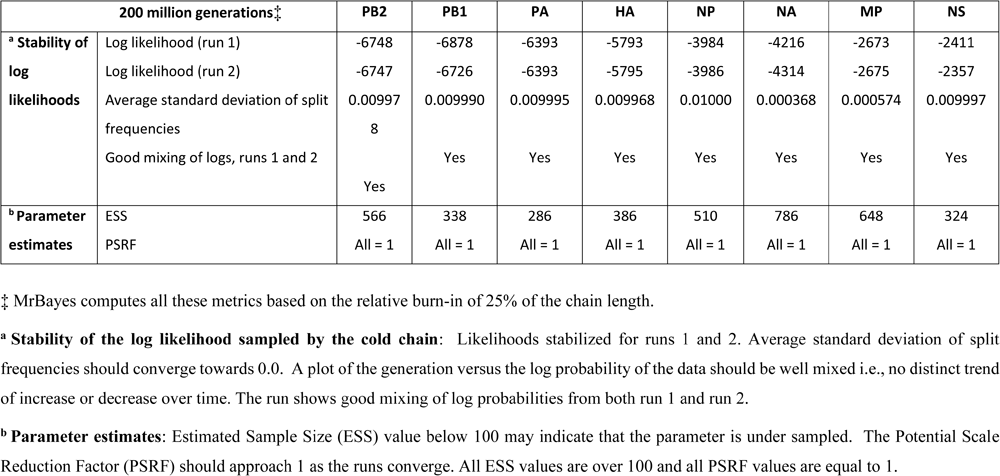
MrBayes convergence diagnostics for A(H1N1)pdm09 viruses.

| 200 million generations‡ |  | PB2 | PB1 | PA | HA | NP | NA | MP | NS |
| --- | --- | --- | --- | --- | --- | --- | --- | --- | --- |
| **<sup>a</sup> Stability of log likelihoods** | Log likelihood (run 1) | -6748 | -6878 | -6393 | -5793 | -3984 | -4216 | -2673 | -2411 |
|  | Log likelihood (run 2) | -6747 | -6726 | -6393 | -5795 | -3986 | -4314 | -2675 | -2357 |
|  | Average standard deviation of split frequencies | 0.009978 | 0.009990 | 0.009995 | 0.009968 | 0.01000 | 0.000368 | 0.000574 | 0.009997 |
|  | Good mixing of logs, runs 1 and 2 | Yes | Yes | Yes | Yes | Yes | Yes | Yes | Yes |
| **<sup>b</sup> Parameter estimates** | ESS | 566 | 338 | 286 | 386 | 510 | 786 | 648 | 324 |
|  | PSRF | All = 1 | All = 1 | All = 1 | All = 1 | All = 1 | All = 1 | All = 1 | All = 1 |

‡ MrBayes computes all these metrics based on the relative burn-in of 25% of the chain length.
**<sup>a</sup> Stability of the log likelihood sampled by the cold chain:** Likelihoods stabilized for runs 1 and 2. Average standard deviation of split frequencies should converge towards 0.0. A plot of the generation versus the log probability of the data should be well mixed i.e., no distinct trend of increase or decrease over time. The run shows good mixing of log probabilities from both run 1 and run 2.
**<sup>b</sup> Parameter estimates:** Estimated Sample Size (ESS) value below 100 may indicate that the parameter is under sampled. The Potential Scale Reduction Factor (PSRF) should approach 1 as the runs converge. All ESS values are over 100 and all PSRF values are equal to 1.

**Table 2.**
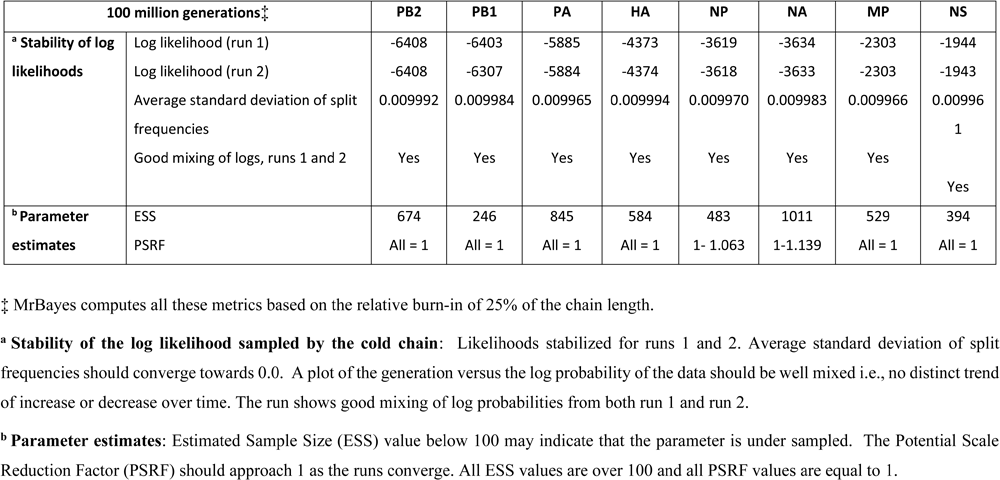
MrBayes convergence diagnostics for A(H3N2) viruses.

### Spatial dynamics of spread of IAVs

We conducted a phylogeographical analysis to assess the global spread of A(H1N1)pdm09 and A(H3N2) viruses using methods implemented in BEAST v1.10.4 package [44], with an asymmetric discrete trait approach that applied the Bayesian stochastic search variable selection (BSSVS) model. To reduce the complexity of the maximum clade credibility (MCC) inference, we categorized location states into geographical regions (“Africa”, “Asia”, “E-SE Asia”, “Europe”, “North America”, “South America”, or “Oceania”) and visualized phylogeographic inferences with the SPREAD3 software v0.9.7.1c package [45]. Due to the important role of E-SE Asia in the global spread of influenza viruses, we partitioned Asia into E-SE Asia and rest of Asia to elucidate the geographical transition states associated with E-SE. To visualize the geographic spread of the virus over time, we generated a D3 file using SPREAD3 v0.9.7.1c package and used a global geo.json file for visualization. We then used the resulting log files to calculate the estimated virus migration rates between regions and Bayes factor (BF) values for significant migration rates between discrete locations: we deemed ≥1,000 as decisive support, 100≤BF<1000 as very strong support, 10≤BF<100 as strong support, and 3≤BF<10 as supported rates.

We also used the ML tree topologies for A(H1N1)pdm09 and A(H3N2) viruses from our mugration models to estimate the number of virus transmission events between sub-Saharan Africa and the rest of the world. Using the date and location-annotated tree topologies, we counted the number of transitions within and between sub-Saharan Africa and the rest of the world and plotted using ggplot2 v3.3.3 R package [37].

### Ethics

Scientific and ethical clearance for the study was obtained from institutional ethics review boards within each study country in Africa, the UK, and USA [26]. Additional ethical approval was sought and received from KEMRI Scientific Steering Committee (SSC# 1055 and 1433) and Oxford Tropical Research Ethics Committee (OxTREC# Ethics ID:60-90). Informed consent was sought and received from the study participants for the study.

## Results

### IAV sequencing and codon-complete genome assembly of IAV genomes

Of the 138 IAV positive samples that were available from the PERCH study, 100 (73%) were successfully sequenced. The recovered genome sequences were classified into A(H1N1)pdm09 (n=31) and A(H3N2) (n=69). These were combined with the additional sequences from Kenya (106 for A(H1N1)pdm09 and 16 for A(H3N2)). Among A(H1N1)pdm09 viruses from the PERCH study, all viruses from 2011-13 fell into six clades namely, clade 3 (n=5), clade 7 (n=1), clade 8 (n=2), clade 6A (n=1), clade 6B (n=2), and clade 6C (n=20), **Figure 1** and **Figure S1B**. Among A(H1N1)pdm09 viruses from the countrywide Kenya study, all viruses from 2011-13 fell into clade 6 (n=96) and clade 6C (n=10), **Figure 1** and **Figure S1B**. For A(H3N2) viruses, all PERCH Africa study viruses from 2011-13 fell into five clades namely, clade 3A (n=6), clade 3B (n=2), clade 3C.2 (n=1), clade 3C.3 (n=39), and clade 7 (n=21), **Figure 2** and **Figure S1C**. Among A(H3N2) viruses from the KCH paediatric study, all viruses from 2011-13 fell into clade 3B (n=4) and clade 7 (n=12), **Figure 2** and **Figure S1C**.

**Figure 1.**
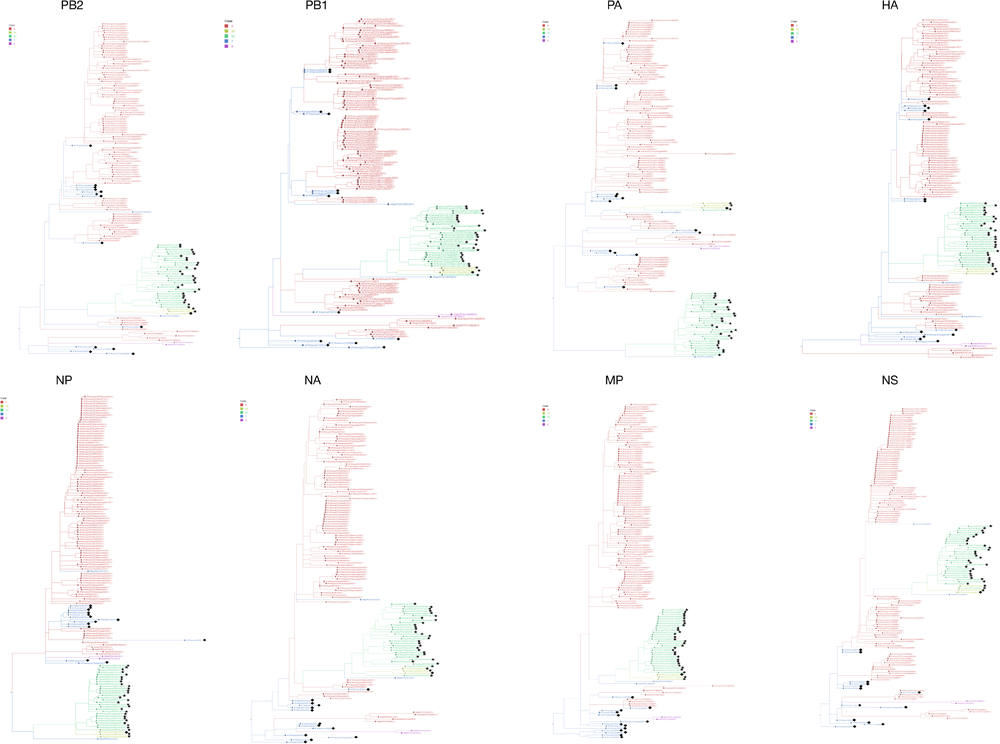
Phylogenetic trees of the eight individual gene segments of influenza A(H1N1)pdm09 viruses from sub-Saharan Africa from the 2011-13 influenza seasons. The tree branches and tips are coloured by virus clade, as shown on the colour-coded key. The positions of intra-subtype reassortant viruses in each phylogeny are indicated by filled stars for PERCH study viruses while viruses from other sub-Saharan African countries are indicated by filled diamonds. PERCH, Pneumonia Etiology Research for Child Health.

**Figure 2.**
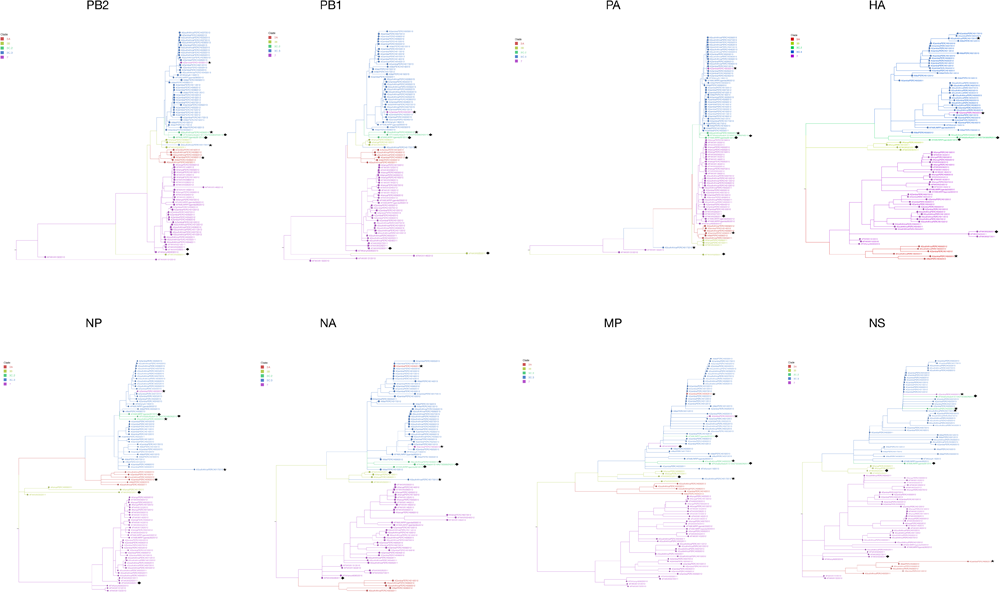
Phylogenetic trees of the eight individual gene segments of influenza A(H3N2) viruses from sub-Saharan Africa from the 2011-13 influenza seasons. The tree branches and tips are coloured by virus clade, as shown on the colour-coded key. The positions of intra-subtype reassortant viruses in each phylogeny are indicated by filled stars for PERCH study viruses while viruses from other sub-Saharan African countries are indicated by filled diamonds. PERCH, Pneumonia Etiology Research for Child Health.

### Patterns of reassortment of IAVs from sub-Saharan Africa

We investigated the reassortment patterns by comparing the position of viruses on the eight segment-specific phylogenies of A(H1N1)pdm09 and A(H3N2) viruses to identify inconsistencies arising from intra-subtype reassortment. For most of the virus strains, those belonging to the same clade, as inferred from HA phylogeny, clustered together on phylogenies generated from the seven remaining gene segments for A(H1N1)pdm09 (**Figure 1**) and A(H3N2) viruses (**Figure 2**), respectively. However, in 44 (29%) A(H1N1)pdm09 virus (**Figure 1**) and eight (9%) A(H3N2) virus (**Figure 2**), intra-subtype reassortants were suspected based on tip locations of viruses in the gene segment phylogenies. Of these reassortants, 31 A(H1N1)pdm09 and four A(H3N2) viruses were identified among our PERCH study samples. GiRaF correctly identified all the 52 intra-subtype reassortant viruses. Interestingly, ten of the 44 A(H1N1)pdm09 and two of the eight A(H3N2) reassortant viruses were recently identified in a continentwide reassortment analysis of IAVs in Africa using GiRaF [46].

The relatedness of these reassortant viruses were visualized using tanglegrams based on pairwise phylogenetic congruence between codon-complete gene segments for A(H1N1)pdm09 (**Figure 3**) and A(H3N2) (**Figure 4**) viruses, with tanglegram twines colored by virus clade to highlight reassortant strains. Additionally, for the viruses detected in sub-Saharan Africa, coalescent model analysis estimated a mean reassortment rate of 0.0225 [95% HPD, 0.1272-0.3231] events/lineage/year among A(H1N1)pdm09 viruses and a mean reassortment rate of 0.0577 [95% HPD, 0.0146-0.1027] events/lineage/year among A(H3N2) viruses. We further observed that the reassortant A(H1N1)pdm09 and A(H3N2) viruses persisted in circulation for 1-2 consecutive years in sub-Saharan Africa based on their date-annotated tree topologies.

**Figure 3.**
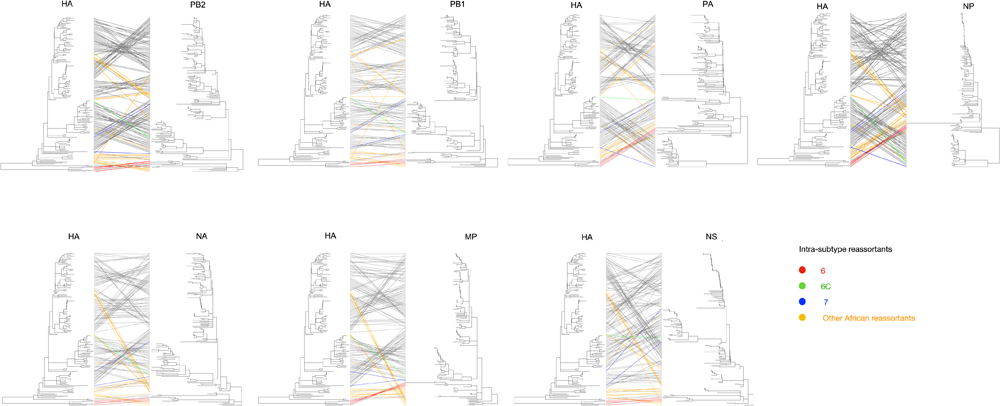
Evolutionary relationships of each gene segment of influenza A(H1N1)pdm09 viruses from sub-Saharan Africa, displaying intra-subtype reassortment with the subtype, highlighting the reassortant strains by clade from the 2011-13 influenza seasons.

**Figure 4.**
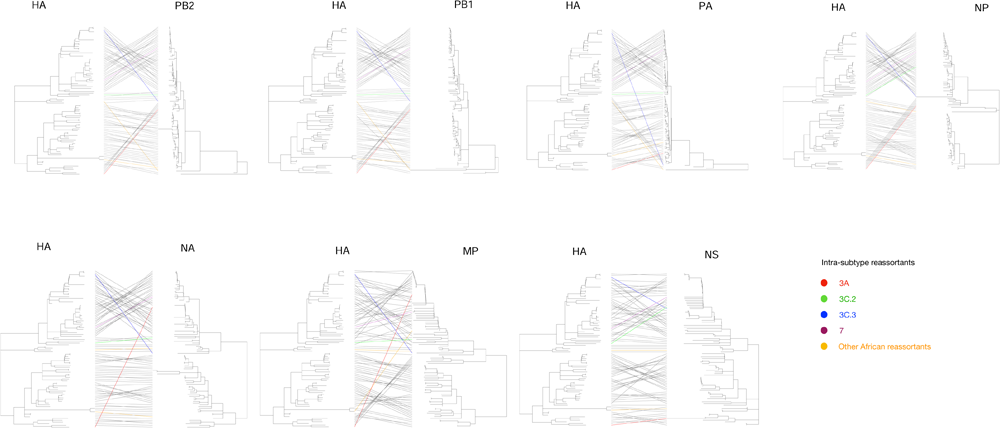
Evolutionary relationships of each gene segment of influenza A(H3N2) viruses from sub-Saharan Africa, displaying intra-subtype reassortment with the subtype, highlighting the reassortant strains by clade from the 2011-13 influenza seasons.

### Viral imports and exports in sub-Saharan Africa

We assessed how 150 A(H1N1)pdm09 virus and 94 A(H3N2) virus genomes from sub-Saharan Africa compared to 441 A(H1N1)pdm09 and 749 A(H3N2) contemporaneous virus genomes, sampled globally by inferring their phylogenies. For A(H1N1)pdm09 virus, we inferred 15 importations originating from outside sub-Saharan Africa (seven from North America, six from Asia, and two from Europe), which represent 15 independent introductions into sub-Saharan Africa from geographical locations outside the region, **Figure 5A**. We also inferred four virus location transition events among the sub-Saharan African countries. Additionally, we captured six virus export events from sub-Saharan Africa to North America, Asia, and Europe, all occurring from Kenya, **Figure 5A**. For A(H3N2) viruses, we inferred 10 importations originating from outside sub-Saharan Africa (three each from North America and Asia and two each from Oceania and South America), which represent 10 independent introductions into sub-Saharan Africa from geographical locations outside the region, **Figure 5B**. Furthermore, we inferred 13 virus location transition events among the sub-Saharan African countries. Additionally, six virus export events from sub-Saharan Africa to North America and Oceania were inferred (**Figure 5B**).

**Figure 5.**
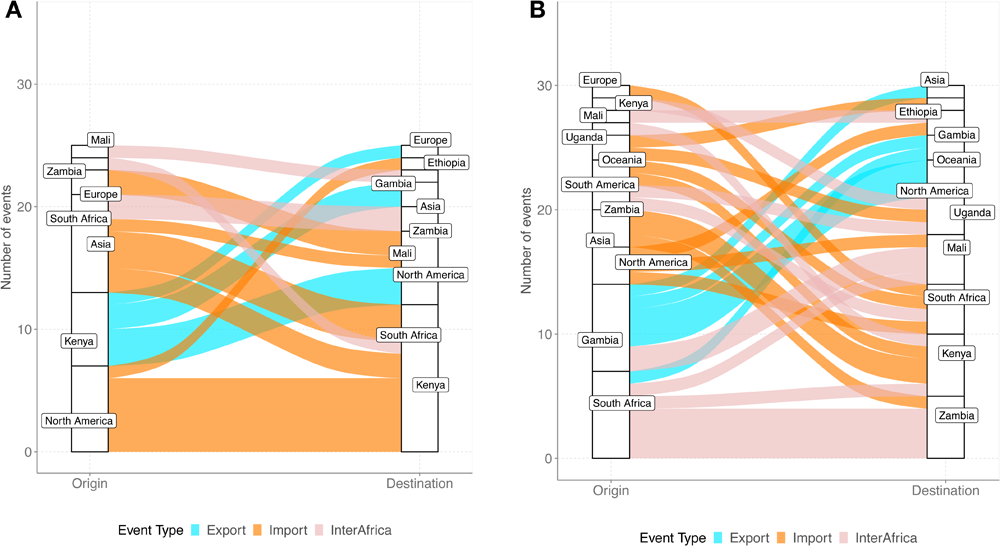
Number of viral imports and exports from sub-Saharan Africa shown as an alluvium plot using virus sequences from sub-Saharan African countries, Asia, Europe, North America, South America, and Oceania for (**A**) influenza A(H1N1)pdm09 viruses and (**B**) influenza A(H3N2) viruses.

### Global migration dynamics of IAVs

For A(H1N1)pdm09 virus, we observed significant migration pathways from E-SE Asia (0.81-3) into multiple geographical regions including sub-Saharan Africa, Asia, Europe, North America, and Oceania. Additionally, we observed significant migration pathways from North America (0.52-0.73) to E-SE Asia, Europe, and Oceania, **Table 3**. The observed migration pathways corroborate these results, **Figure S2**. For A(H3N2) virus, we observed significant migration pathways from E-SE Asia (0.62-3.34) into all other geographical regions including sub-Saharan Africa, and from North America (0.6-1.63) to Europe, Oceania, and South America, **Table 4**. The global migration pathways for A(H3N2) corroborate these results, **Figure S3**.

**Table 3.** Asymmetrical migration rates between global location states inferred using the BSSVS model for influenza A(H1N1)pdm09 virus. BSSVS; Bayesian stochastic search variable selection (BSSVS) model.

| Migration rates* |  |  |  |  |  |  |  |
| --- | --- | --- | --- | --- | --- | --- | --- |
| † | Africa | Asia | E-SE Asia | Europe | North America | Oceania | South America |
| Africa | — | 0.97 | 0.95 | 0.97 | 0.98 | 0.96 | 0.99 |
| Asia | 0.96 | — | **0.88** | 0.83 | 0.99 | 0.95 | 0.95 |
| E-SE Asia | **1.48** | **0.81** | — | **2.83** | **3** | **1.5** | 0.89 |
| Europe | 0.93 | 0.94 | 0.95 | — | 0.95 | 0.95 | **0.56** |
| North America | 0.87 | 1.01 | **0.52** | **0.73** | — | **0.58** | 0.95 |
| Oceania | 0.84 | 0.88 | 0.86 | 0.81 | 0.87 | — | 0.88 |
| South America | 0.82 | 0.85 | 0.84 | 0.93 | 0.84 | 0.88 | — |

\*Migration rates in bold indicate supported rates with Bayes factor $\geq 3$ .
†Number of sequences: sub-Saharan Africa, 150; Asia, 17; E-SE Asia, 131; Europe, 82; North America, 170; South America, 2; and Oceania, 39.

**Table 4.** Asymmetrical migration rates between global location states inferred using the BSSVS model for influenza A(H3N2) virus. Bayesian stochastic search variable selection (BSSVS) model.

| † | Migration rates* |  |  |  |  |  |  |
| --- | --- | --- | --- | --- | --- | --- | --- |
|  | Africa | Asia | E-SE Asia | Europe | North America | Oceania | South America |
| Africa | — | 0.98 | 0.97 | 0.99 | 0.69 | 0.79 | 0.97 |
| Asia | 0.91 | — | 0.92 | 0.92 | 0.96 | 0.96 | 0.93 |
| E-SE Asia | **0.77** | **0.92** | — | **1.19** | **2.27** | **3.34** | **0.62** |
| Europe | 0.93 | 0.97 | 0.97 | — | 0.9 | 0.96 | 0.93 |
| North America | 0.96 | 0.97 | 0.85 | **0.6** | — | **1.63** | **0.93** |
| Oceania | 0.87 | 0.94 | **0.78** | **0.8** | 0.98 | — | 0.98 |
| South America | 0.82 | 0.92 | 0.79 | 0.89 | 0.72 | 0.91 | — |

\*Migration rates in bold indicate supported rates with Bayes factor $\geq 3$ .
†Number of sequences: sub-Saharan Africa, 94; Asia, 17; E-SE Asia, 179; Europe, 38; North America, 178; South America, 199; and Oceania, 138.

## Discussion

We observed that IAV strains from 2011-13 from our study evolved over consecutive influenza seasons and fell into distinct A(H1N1)pdm09 and A(H3N2) virus clades, some of which persisted in circulation as intra-subtype reassortants for consecutive influenza seasons. Furthermore, we observed multiple introductions of IAVs into sub-Saharan Africa over consecutive influenza seasons, with viral importations originating from multiple global geographical regions, for example, North America, Asia, Europe, and Oceania. Additionally, we inferred virus transfer between the sub-Saharan African countries and virus exports from sub-Saharan Africa to other geographical regions, for example, North America, Asia, and Europe. On a global scale, IAVs spread from multiple geographical regions to multiple geographical destinations including sub-Saharan Africa.

Co-circulation of different IAV subtypes and clades is common during epidemics of seasonal influenza, which may facilitate intra-subtype reassortment events among circulating viruses [47–51]. Overall, the reassortment rates among the sub-Saharan African A(H1N1)pdm09 viruses (0.1272-0.3231) were comparable to the 0.15-0.8 events/lineage/year estimated among global A(H1N1)pdm09 viruses [41]. However, sub-Saharan African A(H3N2) viruses reassorted at a lower mean rate of 0.0577 [95 per cent HPD, 0.0146-0.1027] than the global estimate of 0.35-0.65 events/lineage/year [41]. Although A(H3N2) viruses undergo frequent reassortment events globally, the low reassortment rate among some A(H3N2) viruses might be due to the resulting reassortant viruses being unfit and negatively selected, thus not detected in appreciable frequencies [52, 53]. Circulation of intra-subtype reassortants in our study for 1-2 consecutive years demonstrates that IAVs can persist in the population and spread geographically. A recent continentwide study of the rates and patterns of reassortment of IAVs in Africa revealed that novel reassortant viruses emerged every year, with some reassortant viruses persisting in different countries and regions for 1-5 consecutive years [46]. Persistence of intra-subtype reassortants has been demonstrated previously [52, 54], although the factors associated with such persistence patterns at a population level require further investigation. For example, a reassortant A(H3N2) virus subclade, designated 3C.3A2/re, emerged in 2016-17 influenza season and resulted in more hospitalizations and deaths in the 2017-18 North American influenza season [52]. Increased whole genome sequencing over consecutive influenza seasons, particularly in understudied sub-Saharan Africa regions, would allow for an improved understanding of the frequency and timing of such intra-subtype reassortments and the contribution to the evolutionary and transmission dynamics of seasonal influenza.

Globally, we observed significant migration pathways for IAVs. Taken together, these results suggest that the global spread of A(H1N1)pdm09 and A(H3N2) virus epidemics are driven by different geographical regions, which also includes sub-Saharan Africa, in which E-SE Asia and North America are major transmission sources [17, 19, 20, 55]. Our findings support the notion that influenza viruses persist as temporally migrating metapopulations in which new virus strains can emerge in any geographical region, with the location of the source population changing regularly [17]. This underscores the need for improved influenza surveillance particularly in understudied sub-Saharan Africa regions for complete understanding of the global patterns of spread of influenza.

The paucity of sequence data from other African countries limited our analysis of the patterns of spread and persistence of IAVs in the continent, which might have been useful to demonstrate intra-continental spread of influenza viruses in detail, since persistence may be facilitated by climatic variability that generates temporally overlapping epidemics in neighboring countries. Despite this paucity, our study leveraged on existing national surveillance studies, for example, from the countrywide surveillance of influenza viruses in Kenya where A(H1N1)pdm09 virus sequences were included. It is possible that Africa plays a bigger role in the spread of influenza, which we might not have captured. Furthermore, the analysis in this report only involved the coding regions of the A(H1N1)pdm09 and A(H3N2) virus gene segments. Although noncoding regions are conserved, mutations that affect viral replication may occur, and this information may not have been captured in this study.

In conclusion, despite sparse data from influenza surveillance in sub-Saharan Africa, our findings support the notion that influenza viruses persist as temporally structured migrating metapopulations in which new virus strains can emerge in any geographical region, including in sub-Saharan Africa; these lineages may have been capable of dissemination to other continents through a globally migrating virus population. Further knowledge of the viral lineages that circulate within understudied sub-Saharan Africa regions is required to inform vaccination strategies in those regions.

## Data Availability

All generated sequence data were deposited in the Global Initiative on Sharing All Influenza Data (GISAID) EpiFluTM database (https://platform.gisaid.org/epi3/cfrontend) under the accession numbers EPI_ISL_509524-EPI_ISL_509526, EPI_ISL_509564-EPI_ISL_509566, EPI_ISL_509655- EPI_ISL_509669, EPI_ISL_509687, EPI_ISL_510040-EPI_ISL_510043, EPI_ISL_510078- EPI_ISL_510080, EPI_ISL_510102, EPI_ISL_510152-EPI_ISL_510159, EPI_ISL_509025-EPI_ISL_509058, EPI_ISL_509396-EPI_ISL_509411, and EPI_ISL_511774-EPI_ISL_511804

## Acknowledgements.

We would like to acknowledge the Pneumonia Etiology Research for Child Health (PERCH) Study Group for their role in the PERCH study design, coordination, implementation, and reporting.

## Supplementary Figures

**Figure S1.**
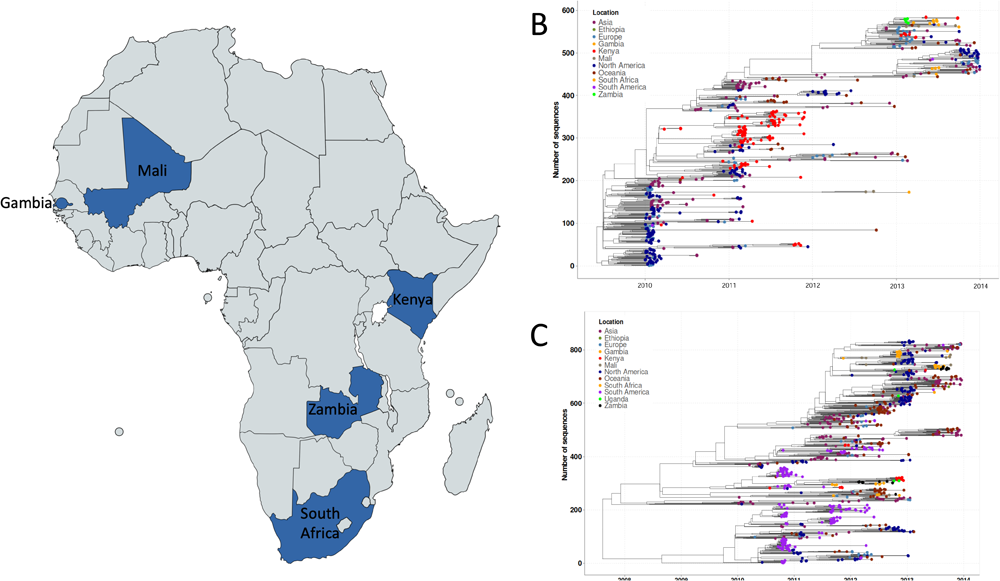
**(A)** PERCH study surveillance sites in sub-Saharan Africa, which include The Gambia, Kenya, Mali, South Africa, and Zambia. PERCH, Pneumonia Etiology Research for Child Health. **(B)** Time-resolved maximum-likelihood phylogenetic tree of influenza A(H1N1)pdm09 viruses from sub-Saharan Africa and other global regions. (**C**) Time-resolved maximum-likelihood phylogenetic tree of influenza A(H3N2) viruses from sub-Saharan Africa and other global regions. The tree tips for figures **B** and **C** are coloured by sampling location, as shown on the colour-coded key.

**Figure S2.**
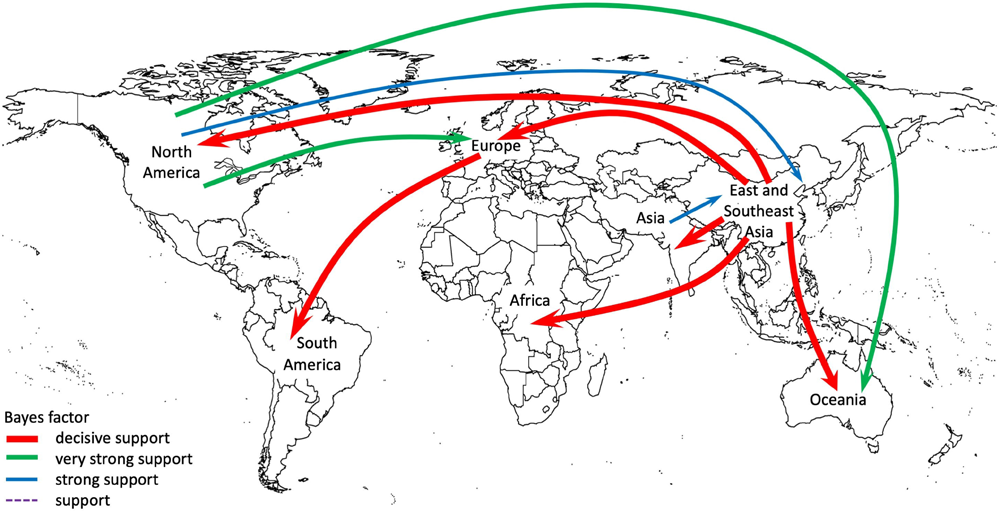
Global dynamics of spread of A(H1N1)pdm09 viruses reconstructed using global virus sequences from sub-Saharan Africa and other global countries. Asymmetric migration pathways between location states were inferred for all global regions. Coloured line arrows indicate significant migration routes from one continent to another, while line thickness represents the degree of statistical support. Red arrowed lines are shown to indicate decisive migration routes with Bayes factor (BF) support ≥1000; green lines represent very strongly supported routes with 100≤BF<1000; blue lines indicate strongly supported routes 10≤BF<100; and purple dotted lines indicate supported routes with 3≤BF<10.

**Figure S3.**
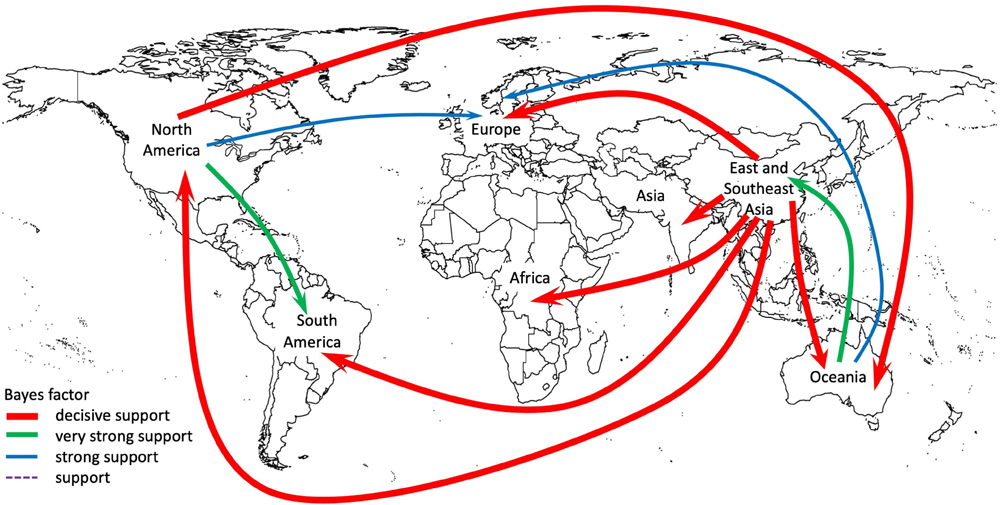
Global dynamics of spread of A(H3N2) viruses reconstructed using global virus sequences from sub-Saharan Africa and other global countries. Asymmetric migration pathways between location states were inferred for all global regions. Coloured line arrows indicate significant migration routes from one continent to another, while line thickness represents the degree of statistical support. Red arrowed lines are shown to indicate decisive migration routes with Bayes factor (BF) support ≥1000; green lines represent very strongly supported routes with 100≤BF<1000; blue lines indicate strongly supported routes 10≤BF<100; and purple dotted lines indicate supported routes with 3≤BF<10.

